# An analysis of the onset and clinical characteristics in cystic lymphatic malformations

**DOI:** 10.1101/2023.09.13.23295470

**Authors:** Naoki Hashizume, Akihiro Fujino, Masataka Takahashi, Ryoya Furugane, Tamotsu Kobayashi, Teizaburo Mori, Eiichiro Watanabe, Motohiro Kano, Akihiro Yoneda, Yutaka Kanamori

## Abstract

**Background:** This study aimed to investigate and compare the applied treatments and the prognosis between patients with congenital and acquired onset cystic lymphatic malformation (cLM).

**Methods:** This study was a retrospective cohort study in a single hospital. Patients with cLM treated at our institution were included in this study, and divided into those diagnosed in utero or at birth (CO) and those diagnosed later after birth (AO) with the acquired emerged lesion. Age, site, and size of the lesion, histological subtype, invasiveness grade of the treatment, and the treatment outcomes were compared.

**Results:** In total, 147 patients were included. The CO group (n = 85) received more invasive treatment, with the distribution of treatment grades differing significantly from the AO group (n = 62, *p* = 0.005). cLMs were mainly located on the neck (n = 53, 36.1%), followed by the head and face (n = 31, 21.1%), axilla (n = 7, 4.8%), chest wall (n = 18, 12.2%), mediastinum (n = 5, 3.4%), abdominal wall (n = 4, 2.7%), retroperitoneum (n = 4, 2.7%), hip (n = 5, 3.4%), arm and hand (n = 8, 5.4%), and leg and foot (n = 12, 8.2%). No significant between-group difference was found in terms of the distribution of the main lesions (*p* = 0.45).

Macrocystic LM was more frequent in the AO group, with a significantly different subtype ratio (*p* = 0.02). The frequency of lower-grade treatment was significantly higher in the macrocystic LM subtype in the AO group (*p* < 0.001), and this group had a significantly higher treatment completion rate (*p* = 0.004). During the treatment course, improvement was seen in 72.9% and 71.0% of patients in the CO group and the AO group, respectively, indicating no significant difference (p=0.79).

**Conclusions:** cLMs that newly appeared after birth tended to be macrocystic and had better outcomes with less-invasive treatment than congenital-onset cLMs. The timing of cLM onset could be a key prognostic factor.

## INTRODUCTION

Common or cystic lymphatic malformation (cLM) is a mass-forming congenital malformation consisting of cystically dilated lymphatic channels^1^. cLM can occur anywhere in the body except in the central nervous system, causing disfigurement, functional impairment, and discomfort. Although cLM is a congenital malformation, approximately 50% of cLM cases are detected after birth, 80%–90% are detected by age 2, and the remainder are detected later ^2–4^.

Major treatment options include course observation ^3–5^, sclerotherapy ^6–9^, surgical resection and, more recently, sirolimus^10–12^ or herbal medicine^13,14^. Spontaneous regression has also been observed^4–7^. cLM lesions vary in terms of location, distribution, tissue subtypes, and symptom severity; therefore, optimal treatment for cLM is frequently challenging to determine, and predicting the final prognosis of lesions post-treatment is even more challenging.

cLM swelling is known to vary according to the amount of lymphatic fluid inside a lesion, ranging from an imperceptible small bulge to a firmly distended large mass often observed in infected lesions or those involving intra-cystic bleeding. In patients with congenitally enlarged lesions that are apparent at birth, the covering skin and blood vessels or the nerves inside the lesion are often permanently distended. This distension remains even after the lesion has shrunk spontaneously or in response to therapy. However, in patients with lesions that have grown and appear later after birth, the surface skin distension, blood vessel, or nerve deviation is often plastic and can return to its original pre-enlargement condition. Based on these observations, we hypothesized that the prognosis may differ between patients with preexisting prenatal lesions and patients who develop lesions after birth and that treatment strategies may need to differ accordingly.

To date, no studies have compared characteristics between patients diagnosed with preexisting prenatal cLM lesions or lesions identified immediately after birth and patients diagnosed with newly emerged lesions after birth. In this study, we aimed to determine differences in applied treatments and outcomes between patients with congenital cLM and those with acquired-onset cLM.

## RESULTS

### Patient demographics and grouping

We reviewed 325 patients with vascular malformations who had been treated in our pediatric surgery department. Of these, 177 patients with cLM were selected. Subsequently, 30 patients with incomplete medical records detailing the onset of treatment or who had visited the department only for a second opinion were excluded. Finally, 147 study patients with cLM were included **(Fig.1).**

**Fig. 1.**
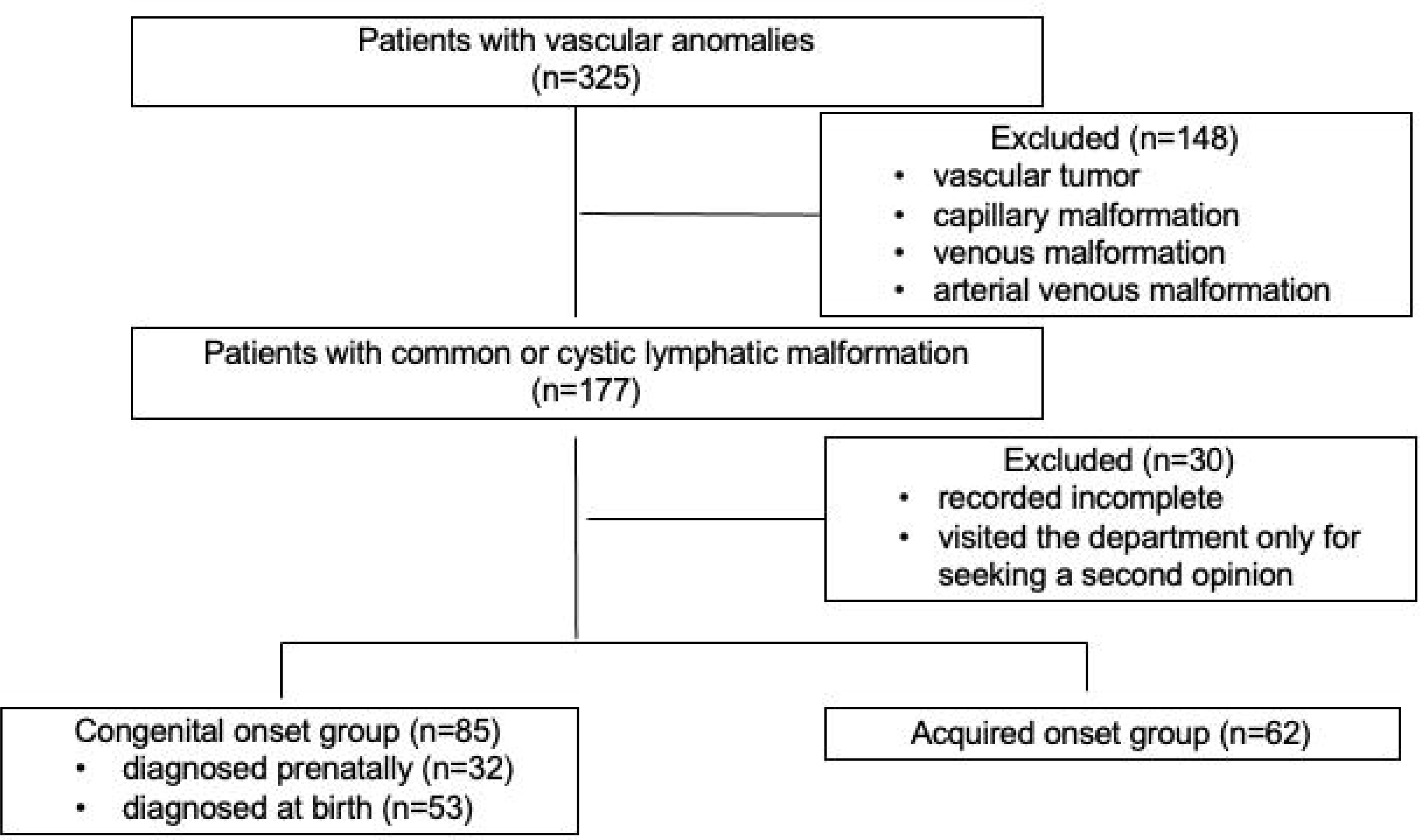
Patients’ flow chart.

The median age of the study population was 70.2 (34.5–100.0) months (**Table 1**), and 71 (48.3%) patients were male. Patients were divided into two groups according to the timing of onset. A congenital-onset (CO) group comprised patients diagnosed with a preexisting lesion, either diagnosed prenatally or immediately after birth. An acquired-onset (AO) group comprised patients diagnosed with a newly appearing lesion after birth.

**Table 1:** Demography of patients and main treatment, and grouping according to the onset timing.

|  | All patients | CO group | AO group | *p*<sup>*</sup> |
| --- | --- | --- | --- | --- |
| **Number** | 147 | 85 | 62 |  |
| **Age (month)** | 70.2 [34.5, 100.0] | 60.9 [32.5, 86.6] | 78.2 [35.2, 115.4] | **0.03** |
| **Sex (male)** | 71 (48.3) | 47 (55.3) | 24 (38.7) | 0.07 |
| **Treatment grade** |  |  |  |  |
| 0 | 55 (37.4) | 23 (27.1) | 32 (51.6) |  |
| 1 | 6 (4.0) | 4 (4.7) | 2 (3.2) |  |
| 2 | 54 (36.7) | 32 (37.7) | 22 (35.4) | **0.005** |
| 3 | 26 (17.7) | 22 (25.9) | 4 (6.5) |  |
| 4 | 6 (4.0) | 4 (4.7) | 2 (3.2) |  |

Age is shown in median [25th, 75th percentile].
Other values are shown in number (percentage).
□ chi-square tests for categorical data comparing CO and AO group. Statistical significance set at $p \leq 0.05$
$p \leq 0.05$ shown in bold

The number of patients with cLM diagnosed prenatally and at birth was 32 (21.8%) and 53 (36.1%), respectively, comprising the CO group. The AO group comprised 62 (42.1%) patients diagnosed one month after birth (**Table 1**).

In the AO group, 25 (40.3%) patients had been diagnosed within the first 12 months of age, 11 (17.7%) at 1 year old, 9 (14.5 %) at 2 years old, 3 (4.8%) at 3 years old, 4 (6.4 %) at 4 years old, and 10 (16.1%) patients had been diagnosed at >5 years old.

The median age of patients in the CO group was significantly lower than that in the AO group (60.9 [32.5–86.6] vs. 78.2 [35.2–115.4] months, respectively, *p* = 0.031). No significant difference was noted between the two groups in terms of sex ratios (*p =* 0.07).

### Demography of the lesions

cLMs were mainly located on the neck (n = 53, 36.1%), followed by the head and face (n = 31, 21.1%), axilla (n = 7, 4.8%), chest wall (n = 18, 12.2%), mediastinum (n = 5, 3.4%), abdominal wall (n = 4, 2.7%), retroperitoneum (n = 4, 2.7%), hip (n = 5, 3.4%), arm and hand (n = 8, 5.4%), and leg and foot (n = 12, 8.2%) (**Table 2**). No significant between-group difference was found in terms of the distribution of the main lesions (*p* = 0.45).

**Table 2:** Demography of the cLM lesions.

|  | All patients | CO group | AO group | *p*<sup>‡</sup> |
| --- | --- | --- | --- | --- |
| **Number** | 147 | 85 | 62 |  |
| **Main location** |  |  |  |  |
| Neck | 53 (36.1) | 35 (41.2) | 18 (29.0) | 0.45 |
| Head and face | 31 (21.1) | 19 (22.4) | 12 (19.4) |  |
| Axilla | 7 (4.8) | 3 (3.5) | 4 (6.5) |  |
| Chest wall | 18 (12.2) | 8 (9.4) | 10 (16.1) |  |
| Mediastinum | 5 (3.4) | 4 (4.7) | 1 (1.6) |  |
| Abdominal wall | 4 (2.7) | 1 (1.2) | 3 (4.8) |  |
| Intra- and retroperitoneum | 4 (2.7) | 1 (1.2) | 3 (4.8) |  |
| Hip | 5 (3.4) | 2 (2.4) | 3 (4.8) |  |
| Arm / Hand | 8 (5.4) | 5 (5.9) | 3 (4.8) |  |
| Leg / Foot | 12 (8.2) | 7 (8.2) | 5 (8.1) |  |
| **Diameter (mm) ‡** | 40 [20, 60] | 35 [20, 50] | 46.5 [23.4, 65] | 0.18 |
| Number | 111 (75.5) | 61 (71.8) | 50 (80.6) |  |
| **Subtype** |  |  |  |  |
| Macrocystic LM | 97 (66.0) | 49 (57.6) | 48 (77.4) | **0.02** |
| Microcystic LM | 37 (25.2) | 25 (29.4) | 12 (19.4) |  |
| Mixed LM | 13 (8.8) | 11 (12.9) | 2 (3.2) |  |

Diameter is shown in median [25th, 75th percentile].
Other values are shown in number (percentage).
‡: Mean of the number of patients in whom the diameter data was recorded.
□ chi-square tests for categorical data comparing CO and AO group. Statistical significance set at $p \leq 0.05$
$p \leq 0.05$ shown in bold

The record of the maximum diameter of each lesion was available for 111 (75.5%) patients (CO group, n = 61 [71.8%]); AO group, n = 50 [80.6%]). In 36 patients, lesion diameter had not been measured when the lesion was located on the tongue or was too large and deemed unsuitable for ultrasound measurement or when there was insufficient cooperation from the patient. The mean maximum diameter was 40 (20–60) mm. No significant difference in diameter was observed between the groups (CO group, 35 [20–50] mm; AO group, 46.5 [23.4–65] mm, *p* = 0.18).

Regarding cLM lesion histological subtypes, 97 (66.0%) were macrocystic, 37 (25.2%) were microcystic, and 13 (8.8%) were mixed type. Macrocystic LM was more frequent in the AO group than in the CO group, with a significant difference in the subtype ratio (*p* = 0.03).

### Treatment grade

Typically, a clinician selects one of the following five treatment grades (grade 0, observation; grade 1, herbal medicine; grade 2, sclerotherapy; grade 3, surgical resection; and grade 4, sirolimus) and balances treatment invasiveness with the anticipated treatment outcome. A higher treatment grade is applied when improvement has not occurred at lower grades. In this study, the treatment grade was defined as the highest grade of treatment during the treatment course. Herbal medicines used by the patients included “Eppikajyututo,” “Ogikenchuto,” and/or “Keishibukuryogankayokuinin,” which are reportedly effective in reducing and eliminating excessive fluids in patients with inflammatory joint disorders and lymph edema^13,14^. Those have diuretic action. For sclerotherapy, OK-432 (Picibanil, Chugai Pharmaceuticals, Tokyo, Japan) was used as a first-line sclerosant and a second-line sclerosant combined with bleomycin.

The number of cases in each treatment grade was as follows: grade 0, n = 55, (37.4%); grade 1, n = 6 (4.0%); grade 2, n = 54 (36.7%); grade 3, n = 26 (17.7%); and grade 4, n = 6 (4.0%) (**Table 1**). The distribution of treatment grades differed significantly between the CO and AO groups (*p* = 0.005).

When classified according to histological subtype, the treatment grade distributions differed significantly between the CO and AO groups in terms of macrocystic LM (*p* = 0.001) but not for microcystic and mixed LM subtypes (*p* = 0.38 and *p* = 0.23, respectively) (**Table 3**).

**Table 3:**
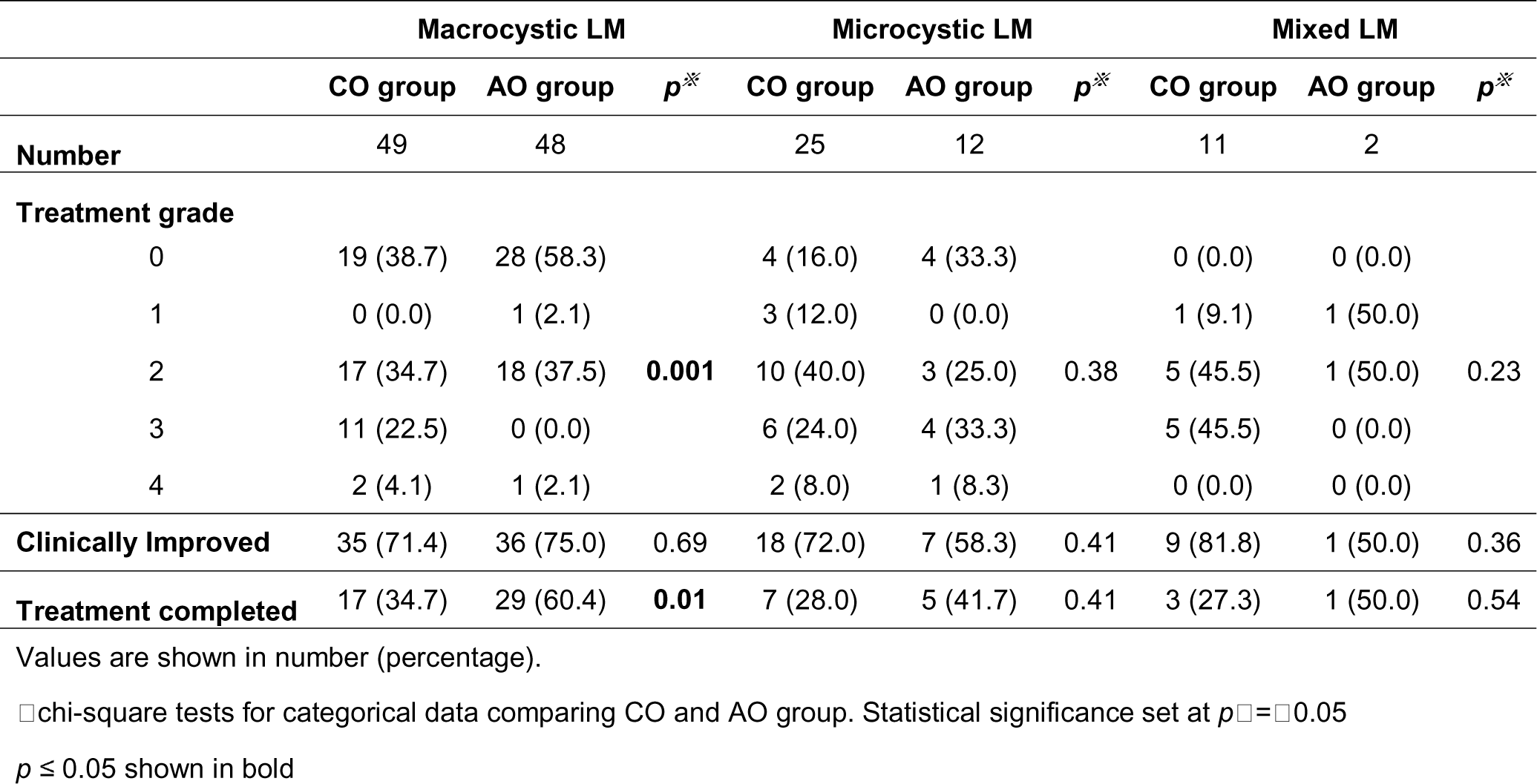
The treatment grade and the outcomes; comparison between subtypes of cLM.

|  | Macrocystic LM |  |  | Microcystic LM |  |  | Mixed LM |  |  |
| --- | --- | --- | --- | --- | --- | --- | --- | --- | --- |
|  | CO group | AO group | *p*<sup>**</sup> | CO group | AO group | *p*<sup>**</sup> | CO group | AO group | *p*<sup>**</sup> |
| **Number** | 49 | 48 |  | 25 | 12 |  | 11 | 2 |  |
| **Treatment grade** |  |  |  |  |  |  |  |  |  |
| 0 | 19 (38.7) | 28 (58.3) |  | 4 (16.0) | 4 (33.3) |  | 0 (0.0) | 0 (0.0) |  |
| 1 | 0 (0.0) | 1 (2.1) |  | 3 (12.0) | 0 (0.0) |  | 1 (9.1) | 1 (50.0) |  |
| 2 | 17 (34.7) | 18 (37.5) | **0.001** | 10 (40.0) | 3 (25.0) | 0.38 | 5 (45.5) | 1 (50.0) | 0.23 |
| 3 | 11 (22.5) | 0 (0.0) |  | 6 (24.0) | 4 (33.3) |  | 5 (45.5) | 0 (0.0) |  |
| 4 | 2 (4.1) | 1 (2.1) |  | 2 (8.0) | 1 (8.3) |  | 0 (0.0) | 0 (0.0) |  |
| **Clinically Improved** | 35 (71.4) | 36 (75.0) | 0.69 | 18 (72.0) | 7 (58.3) | 0.41 | 9 (81.8) | 1 (50.0) | 0.36 |
| **Treatment completed** | 17 (34.7) | 29 (60.4) | **0.01** | 7 (28.0) | 5 (41.7) | 0.41 | 3 (27.3) | 1 (50.0) | 0.54 |

Values are shown in number (percentage).
□ chi-square tests for categorical data comparing CO and AO group. Statistical significance set at $p \leq 0.05$
$p \leq 0.05$ shown in bold

### Treatment outcomes

Post-treatment outcomes improved in 106 (72.1%) of patients who completed or continued to receive treatment. The rate of improvement in the CO and AO groups was 72.9% and 71.0%, respectively, indicating no significant difference (*p* = 0.79) (**Table 4**). There was no significant between-group difference in improvement rates concerning each treatment grade (**Table 5**). When classified according to histological subtype, no significant difference was observed in the improvement rate between the CO and AO groups in terms of macrocystic, microcystic, or mixed LM (*p* = 0.69, 0.41, and 0.36, respectively) (**Table 3**).

**Table 4:** Outcome of the whole treatment.

|  | **All patients** | **CO group** | **AO group** | **$p^{\square}$** |
| --- | --- | --- | --- | --- |
| **Number** | 147 | 85 | 62 |  |
| **Age (month)** | 70.2 [34.5, 100.0] | 60.9 [32.5, 86.6] | 78.2 [35.2, 115.4] | **0.03** |
| **Duration of treatment (month)** | 51.6 [23.4, 80.7] |  | 28.8 [10.5, 72.3] | **0.001** |
| **Improved** | 106 (72.1) | 62 (72.9) | 44 (71.0) | 0.79 |
| **Treatment-completed** |  |  |  |  |
| Number | 62 (42.2) | 27 (31,8) | 35 (56.5) | **0.004** |
| Age (month) | 81.3 [41.8, 122.3] | 67.4 [40.8, 89.9] | 95.5 [56.2, 153.4] | 0.59 |
| Duration of treatment (month) | 55.6 [16.8, 88.6] |  | 32.2 [5.4, 78.4] | **0.02** |

Values are shown in number (percentage) or in median [25th, 75th percentile].
□ chi-square tests for categorical data comparing CO and AO group. Statistical significance set at $p \leq 0.05$
$p \leq 0.05$ shown in bold

**Table 5:** Comparison of the improved rate in each treatment grades.

|  | All patients | CO group | AO group | *p*<sup>※</sup> |
| --- | --- | --- | --- | --- |
| **Number** | 147 | 85 | 62 |  |
| **Treatment grade** |  |  |  |  |
| 0 | 30/55 (54.5) | 11/23 (47.8) | 19/32 (59.4) | 0.40 |
| 1 | 4/6 (66.7) | 3/4 (75.0) | 1/2 (50.0) | 0.55 |
| 2 | 43/54 (79.6) | 23/32 (71.9) | 20/22 (90.9) | 0.08 |
| 3 | 24/26 (92.3) | 21/22 (95.4) | 3/4 (75.0) | 0.23 |
| 4 | 5/6 (83.3) | 4/4 (100) | 1/2 (50.0) | 0.10 |
| **Total** | 106/147 (72.1) | 62/85 (72.9) | 44/62 (71.0) | 0.79 |

Values are Effective cases/ total number (percentage).
□ chi-square tests for categorical data comparing CO and AO group. Statistical significance set at $p \leq 0.05$

### Treatment completion

Treatment was completed successfully for 62 (42.1%) patients, while 85 (57.8%) patients continued receiving treatment or had visited the clinic at the time of the study cut-off date (**Table 4**). The median age of the treatment-completed patients was 81.3 [41.8–122.3] months. In the CO group, 27 (31.8%) patients had completed treatment, whereas 35 (56.5%) patients in the AO group had completed treatment (**Table 4**), and this difference was significant (*p* = 0.004). When classified according to histological subtype, the treatment completion rate was significantly higher in the AO group compared with the CO group for macrocystic LM (*p* = 0.01) but not for microcystic and mixed LM subtypes (*p* = 0.41 and *p* = 0.54, respectively) (**Table 3**).

### Treatment duration

The median treatment duration was 51.6 (23.4–80.7) months (**Table 4**). The median treatment duration in the treatment-completed patients was 55.6 (16.8–88.6) months. The treatment duration in the CO group was significantly longer than in the AO group (60.9 [32.5–86.6] vs. 28.8 [10.5, 72.3] months, respectively; *p* = 0.001). The treatment duration in the treatment-completed patients was significantly longer in the CO group than in the AO group (67.4 [40.8, 89.9] vs. 32.2 [5.4, 78.4] months, respectively; *p* = 0.02).

## DISCUSSION

This study focused on identifying differences in cLM treatment and outcomes between patients in CO and AO groups. We found that cLMs that newly appeared after birth had significantly better outcomes with less-invasive treatment than congenitally apparent cLMs.

The difference between the CO and AO groups was significant in terms of histological subtype distribution, invasiveness grade of treatment, and clinical outcomes. Overall, improvement rates and the effectiveness of each treatment grade were comparable between the AO and CO groups (**Tables 4, 5**), suggesting that treatment selection was reasonable and appropriate. In the AO group, the treatment duration was significantly shorter, and the treatment completion rate was significantly higher (**Table 4**). These results strongly suggest that patients with AO cLM can expect a better treatment process and final prognosis than those with congenital onset.

Concerning the treatment grade, the AO group had more patients under observation (grade 0), and fewer had undergone invasive surgical resection (grade 3) than those in the CO group, indicating less invasive treatment was required for this AO group (**Table 1**) to achieve better outcomes (**Table 4**).

In the AO group, macrocystic LM was significantly more frequent than in the CO group (**Table 2**). This was a reasonable result, given that shrunken macrocystic lesions with low lymphatic fluid content can suddenly distend through the accumulation of intra-cystic lymphatic fluid, intra-cystic hemorrhage, or local infection. It was suspected that the difference in lymph flow in macrocystic LM in AO group patients occurred at the times of onset^4^.

Generally, macrocystic lesions shrink better with treatment than micro- or mixed cystic types. Thus, the AO group was expected to have had better outcomes owing to the higher frequency of macrocystic lesions in that group.

When we compared patients with macrocystic lesions only, the AO group showed a significantly better outcome with treatment completion and with less invasive treatment than the CO group (**Table 3**). Concerning macrocystic LMs in the AO group, no patients underwent surgical resection, approximately 60% of the patients were under observation without surgical intervention, and the treatment-completed rate was approximately 60%. These results can be explained as follows. Patients with congenitally obvious lesions are born with completed deformities of normal structures, such as hyper-stretched skin, separated muscle fibers, or stretched nerves or blood vessels. However, if swelling occurs after birth, healthy tissue tends to retain its plasticity even when swelling results in deformity. Hence, AO lesions, including the macrocystic type, tend to improve more easily with less invasive treatment and return to their original condition before the acquired swelling.

Spontaneous regression has been reported to occur in 12.5–50% of patients with cLM^4–8^. Kato et al. reported that spontaneous regression occurred only in macrocystic or mixed cLMs and in patients aged >2 years at lesion onset^4^. They considered that spontaneous regression of a cLM may be associated with lymph flow pattern variations, with an increased inflow overcoming outflow drainage capacity, thus resulting in temporary expansion^4^.

In this study, 37.4% of patients with cLM remained under observation only (**Table 1**). Clinical improvement was observed in 54.5% of these cases (**Table 5**), suggesting spontaneous regression occurred in approximately 20% of patients with cLM. When comparing histological subtypes, only 16% of patients (CO group, n = 4 of 25; AO group, n = 4 of 12) with microcystic or mixed LM (CO group, n = 0 of 11; AO group, n = 0 of 2) were under observation (**Table 3**), whereas 48.5% of patients (CO group, n = 19 of 49; AO group, n = 28 of 48) with macrocystic LM were under observation. These findings are consistent with those of previous studies^5–8^.

In our study, 27.1% of patients in the CO group and 51.6% in the AO group were under observation (**Table 1**), with improvements in the clinical condition noted in 47.8% and 59.4% of such patients, respectively (**Table 5**). Therefore, the management of patients under observation was effective for 13.0% and 30.7% of the patients in the CO and AO groups, respectively (*p* = 0.01). Thus, we concluded that more patients with cLM in the AO group had undergone spontaneous regression than those in the CO group. Our results support previous findings concerning spontaneous regression and suggest that the timing of onset could influence the clinical outcome.

In this study, Japanese herbal medicine was graded as the second level of treatment after observation. Japanese herbal medicine is often used in children. Some studies have reported that treatment with Japanese herbal medicine has resulted in a marked reduction in cLM^13,14^. There is currently no consensus concerning the administration of Japanese herbal medicine to reduce cLM, as clear evidence is lacking in terms of evaluating its efficacy. However, our department uses Japanese herbal medicine as an alternative treatment for patients with cLM. Six patients were prescribed herbal medicine, which was effective in four cases (data not shown).

Six patients were administered sirolimus in this study. This mTOR inhibitor has been shown to act on lymphatic endothelium in the lesion to suppress proliferation and regulate lymph production and leakage through decreasing lymphatic endothelial cell activity^10–12^. Ozeki et al. reported that 4 of 5 (80%) patients with cLM showed a partial response on magnetic resonance imaging (MRI); moreover, their disease severity and quality of life scores improved significantly^12^. However, its efficacy and safety should be investigated further^11,12^ as the best indication for which type or size of cLM remains unknown. In this study, we set sirolimus at grade 5 because, to date in Japan, sirolimus has only been used for intractable cases in which other treatments have been ineffective. Although the grade definitions may be controversial, we consider that the definitions used in our study did not substantially affect our results.

Treatment duration and completion were significantly shorter in the AO group (**Table 4**). This finding suggests that patients with cLMs in the AO group could expect a better prognosis than those with cLMs in the CO group since treatment duration is a key index of disease severity. We found that the goal of treatment for cLM differed for each patient. For each lesion, balancing between the degree of expected improvement obtained and the degree of treatment invasiveness is essential, and quality of life must also be considered. Therefore, it remains challenging to objectively assess what treatment type is most appropriate and whether a patient would benefit from invasive treatment. Our goal was to assess patient satisfaction and the point of completion of outpatient visits. While patient satisfaction cannot be used to confirm whether a cLM has been treated completely, consideration of this factor serves as a practical indicator of the effectiveness of treatment.

This single-center study had some limitations. First, the sample size was small, considering the wide range of age distribution and various locations and types of lesions. A multicenter study with a larger number of patients with cLM should be performed to verify our study results. Second, we did not determine the endpoint of clinical improvement using numerical and objective values. The volume reduction rate measured using imaging volumetry is generally used to evaluate treatment effectiveness for mass lesions. We recommend an objective examination of treatment outcomes using imaging tools. However, volumetry on imaging studies and body surface evaluations may occasionally differ. Even if the degree of reduction in an image is significant, the appearance sometimes does not change. We considered that termination of treatment and outpatient clinic visits, following an agreement between patients, their families, and the clinician, could reflect positive outcomes more accurately than measuring lesion volume reduction rates. Third, this was a retrospective study. Prospective studies are recommended to verify the accuracy of our results.

In conclusion, our results suggest that cLMs newly appearing after birth tend to be macrocystic and have better outcomes with less invasive treatment than cLMs with lesions that emerge congenitally or at birth. We recommend that pediatric patients with LMs diagnosed later after birth should be observed, especially those with a macrocystic lesion type in the early stages of onset if they do not exhibit severe life-threatening symptoms. Since the two groups showed clear differences in terms of prognoses, future studies of cLM should consider onset-timing when developing a research plan and analysis.

## METHODS

### Diagnosis and inclusion criteria

All patients with vascular malformations and those who underwent pediatric surgery at the National Center for Child Health and Development between January 2016 and October 2021 were reviewed. Patients diagnosed with cLM were selected carefully, excluding other LMs, such as generalized lymphatic anomaly, Gorham–Stout disease, lymphangiectasia, or primary lymphedema^15^. Patients with incomplete medical records concerning onset or treatment and those who had visited our department for a second opinion were excluded.

cLM was diagnosed through physical examination and imaging studies such as ultrasonography, computed tomography, and/or MRI. cLM lesions were classified as macrocystic, microcystic, or mixed, based on ultrasound findings, in accordance with the International Society for the Study of Vascular Anomalies classifications^15^.

### Collected clinical values

The maximum diameter of each lesion was measured during the imaging examinations performed at the time of diagnosis. The main location of the lesion was defined as the area of the body where the cLM initially originated. The main body areas included the head and face, neck, axilla, chest wall, mediastinum, abdominal wall, intra- and retroperitoneum, hip, arm and hand, and leg and foot.

cLMs vary in size and location and can involve surrounding organs, sometimes resulting in life-threatening complications, which needs to be considered when determining an appropriate therapeutic strategy for each patient.

Improvement was defined as improvement in lesion appearance with swelling reduction, which was evaluated on photographs or as a reduction in volume on imaging studies taken at follow-up compared with those taken at treatment initiation. When a lesion had reduced in size and was evaluated as being too small for further treatment, or when a lesion appeared to have improved based on its appearance to a satisfactory level regardless of the actual residual lesion size, treatment was terminated with the patient’s consent. Until a satisfactory appearance was achieved, patients continued to visit the hospital regularly. Treatment completion was defined as sufficient clinical improvement in the lesion and termination of regular hospital visits. The duration of treatment was defined as the period from the time of disease onset to the time of outcome determination (**Fig. 2**).

**Fig. 2.**
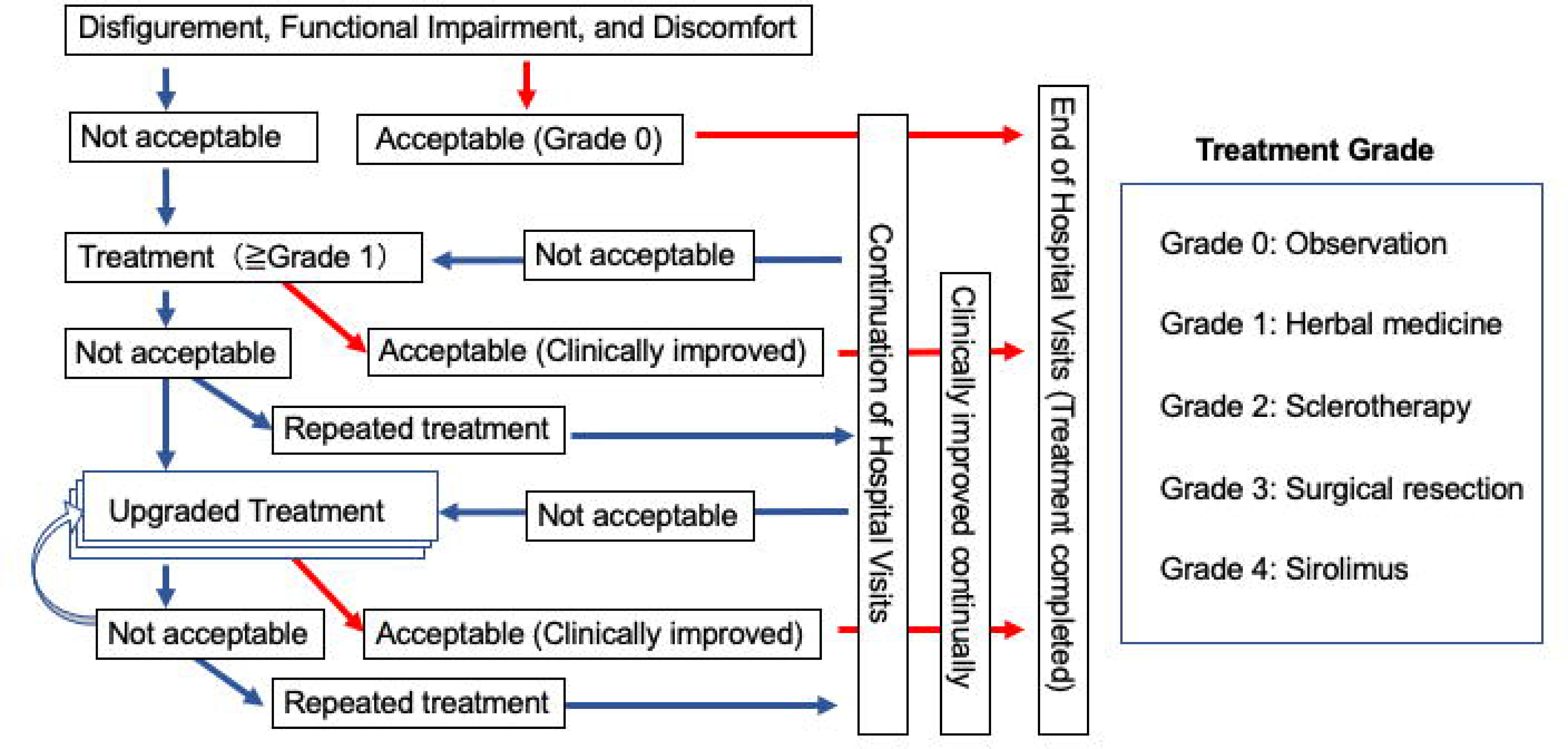
Treatment algorithm.

The cut-off date for recording the outcome for each patient was determined to be either October 31, 2021, or the day of the last visit for patients whose treatment had been completed. Patient age indicated the age at which the outcomes were determined.

### Data analysis

Patient age at onset, tissue subtype, the primary location of the cLM lesion, treatment grade, clinical improvement, age at treatment termination, and clinical outcomes were analyzed retrospectively. Data are presented as median and interquartile ranges. The CO and AO groups were compared using Mann–Whitney U and chi-square tests. Statistical significance was set at *p* < 0.05. All statistical analyses were performed using JMP^®^ version 16.0 (SAS Institute Japan, Tokyo, Japan) software.

## Declarations

## Ethics approval and consent to participate

This study was approved by the Ethics Committee of the National Center for Child Health and Development (Approval No. 2021-833). Informed consent was obtained in the form of an opt-out on the website. Those who rejected were excluded.

## Consent for publication

Not applicable.

## Availability of data and materials

The data that support the findings of this study are available from the corresponding author on reasonable request.

## Competing interests

The authors declare that they have no competing interests.

## Funding

This study was supported by a grant from the Research Project for Intractable Diseases by the Ministry of Health, Labor, and Welfare in Japan (grant no. JPMH20FC1017 and JPMH20FC1042), received by Noriaki Usui (N.U.) and Tomoaki Taguchi (T.T.), respectively, and by a National Center for Child Health and Development grant (2021B-10), received by A.F. N.U and T.T. did not contribute to this study and are not listed as study co-authors.

## Authors’ contributions

F.A. and N.H.: Conceptualization, Design, Writing - Original draft preparation. M.T., R.F. T.K. T.M. E.W. and M.K.: Patient management. A.Y.and Y.K.: Manuscript revision - Critical review for intellectual content. All authors read and approved the final version for publication.

## Abbreviations

cLM: Common or cystic lymphatic malformation
LM: Lymphatic malformation
MRI: Magnetic resonance imaging
CO: Congenital-onset
AO: Aquired-onset

## Acknowledgements

The authors would like to thank Editage (http://www.editage.jp) for English language editing.

## Notes

### Competing Interest Statement

The authors have declared no competing interest.

